# A Comprehensive Study on the Impact of Hypertension on Bone Metabolism Abnormalities Based on NHANES Data and Machine Learning Algorithms

**DOI:** 10.1101/2024.09.07.24313248

**Authors:** Jinyao Li, Mingcong Tang, Ziqi Deng, Yanchen Feng, Xue Dang, Lu Sun, Yunke Zhang, Jianping Yao, Min Zhao, Feixiang Liu

## Abstract

**Background:** Hypertension (HTN), a globally prevalent chronic condition, poses a significant public health challenge. Concurrently, abnormalities in bone metabolism, such as reduced bone mineral density (BMD) and osteoporosis (OP), profoundly affect the quality of life of affected individuals. This study aims to comprehensively investigate the relationship between HTN and bone metabolism abnormalities using data from the National Health and Nutrition Examination Survey (NHANES) and advanced machine learning techniques.

**Methods:** Data were sourced from the NHANES database, covering the years 2009 to 2018. Specifically, femur and spine BMD measurements were obtained via dual-energy X-ray absorptiometry (DXA) for the 2009–2010 period, given the lack of full-body data. A predictive model was developed to estimate total body BMD from femur and spine measurements. The initial dataset comprised 49,693 individuals, and after rigorous data cleaning and exclusion of incomplete records, 7,566 participants were included in the final analysis. Data were processed and analyzed using SPSS, which facilitated descriptive statistical analysis, multivariate logistic regression, and multiple linear regression, alongside subgroup analyses to explore associations across different demographic groups. Machine learning algorithms, including neural networks, decision trees, random forests, and XGBoost, were utilized for cross-validation and hyperparameter optimization. The contribution of each feature to the model output was assessed using SHAP (Shapley Additive Explanations) values, enhancing the model’s accuracy and robustness.

**Results:** Baseline characteristic analysis revealed that compared to the non-HTN group, the HTN group was significantly older (44.37 vs. 34.94 years, p < 0.001), had a higher proportion of males (76.8% vs. 60.7%, p < 0.001), higher BMI (31.21 vs. 27.77, p < 0.001), a higher smoking rate (54.4% vs. 41.2%, p < 0.001), and notably lower BMD (1.1507 vs. 1.1271, p < 0.001). When comparing the low bone mass group with the normal bone mass group, the former was older (36.02 vs. 34.5 years, p < 0.001), had a lower proportion of males (41.8% vs. 63.3%, p < 0.001), lower BMI (25.28 vs. 28.25, p < 0.001), and a higher incidence of HTN (10.9% vs. 8.6%, p = 0.006). Overall logistic and multiple linear regression analyses demonstrated a significant negative correlation between HTN and bone metabolism abnormalities (adjusted model Beta = −0.007, 95% CI: −0.013 to −0.002, p = 0.006). Subgroup analysis revealed a more pronounced association in males (Beta = −0.01, p = 0.004) and in the 40–59 age group (Beta = −0.01, p = 0.012). The machine learning models corroborated these findings, with SHAP value analysis consistently indicating a negative impact of HTN on BMD across various feature controls, thus demonstrating high explanatory power and robustness across different models.

**Conclusion:** This study comprehensively confirms the significant association between HTN and bone metabolism abnormalities, utilizing NHANES data in conjunction with machine learning algorithms.

## Introduction

Hypertension (HTN), as a globally prevalent chronic disease, significantly impacts public health[1]. It not only leads to severe complications such as myocardial infarction and ischemic or hemorrhagic stroke[2,3] but is also closely related to various metabolic diseases, particularly bone metabolism abnormalities[4]. Abnormal bone metabolism includes issues such as reduced bone mineral density (BMD) and osteoporosis (OP). These problems not only significantly reduce patients’ quality of life but also increase the risk of fractures and other related complications[5]. Currently, specific statistics on bone metabolism abnormalities (such as OP) among HTN patients worldwide are scarce. However, some studies have initially revealed a subtle link between HTN and bone metabolism abnormalities. For example, one study found that the probability of OP in HTN patients is 2.69 times that of non-HTN patients[6]. HTN affects the occurrence and development of OP through various mechanisms such as reducing blood flow to bone tissue, affecting calcium metabolism, and promoting inflammatory responses[7]. Specifically, common HTN-associated conditions such as increased calcium excretion and abnormal activation of the renin-angiotensin system may lead to increased bone resorption and reduced bone mass[8]. Additionally, chronic inflammatory responses and oxidative stress under HTN conditions also adversely affect bone cell function, further accelerating the progression of OP[9]. Increasing research focuses on the interrelationship between HTN and bone metabolism abnormalities, attempting to reduce the comorbidity risk through comprehensive management strategies. For example, controlling blood pressure may not only reduce cardiovascular events but also help improve bone health[10]. Antihypertensive drugs such as calcium channel blockers and angiotensin-converting enzyme inhibitors have demonstrated a potential protective effect on BMD in addition to their blood pressure-lowering effects[11]. Therefore, exploring the relationship between HTN and bone metabolism abnormalities is of great significance for disease prevention and treatment[12]. Joint research in the cardiovascular and bone disease fields can better understand the association between HTN and bone metabolism abnormalities, formulate individualized prevention and treatment plans, comprehensively assess patients’ health status, provide more precise and effective management strategies, and reduce the dual risks of cardiovascular disease and OP.

With the development of epidemiological research and biostatistical methods, using large-scale databases for association studies or correlation studies has become possible[13]. The National Health and Nutrition Examination Survey (NHANES) database, conducted by the Centers for Disease Control and Prevention, provides extensive demographic, clinical, and laboratory data[14], making it an ideal data source for large-scale epidemiological research[15]. The NHANES database covers multiple rounds of survey data from 2009 to 2018, providing a solid data foundation for related studies[16]. In modern medical research, machine algorithm models have demonstrated their great potential and advantages[17]. By using a deep learning (DL) model developed with NHANES data, it not only outperforms traditional clinical assessment tools and machine learning (ML) models in osteoporosis (OP) classification but also provides personalized risk assessments and explains the contribution of each feature to the model’s results[18]. Machine learning, as a fusion of computer science and statistics, has been successfully applied in many medical specialties, representing the next wave of advancements in modern healthcare[19,20].

To address the data insufficiency in previous studies and comprehensively understand the impact of HTN on bone metabolism abnormalities, this study used NHANES database data from 2009 to 2018 and further elucidated the association between HTN and bone metabolism abnormalities through machine learning algorithms, providing new scientific evidence and public health policy recommendations for the prevention and treatment of bone metabolism diseases such as OP.

## Materials and Methods

### 1.1 Data Sources

The data used in this study were obtained from the NHANES. The NHANES database is a nationwide cross-sectional survey conducted by the CDC to assess the health and nutritional status of the US population[21]. The NHANES survey protocol received ethical approval from the National Center for Health Statistics Ethics Review Board, and all participants provided written informed consent before the survey[22]. This study’s data were sourced from the NHANES database, covering multiple rounds of survey data from 2009 to 2018.

### 1.2 Study Participants

Adults aged 20 years and above from the NHANES database were included, excluding pregnant women and patients with diseases that might affect bone metabolism, such as thyroid diseases, diabetes, and long-term use of glucocorticoids. In this study, we used NHANES data to weight 7,566 samples (Figure 1), making the weighted sample size 13,655, ensuring the representativeness of the analysis results.

**Figure I.**
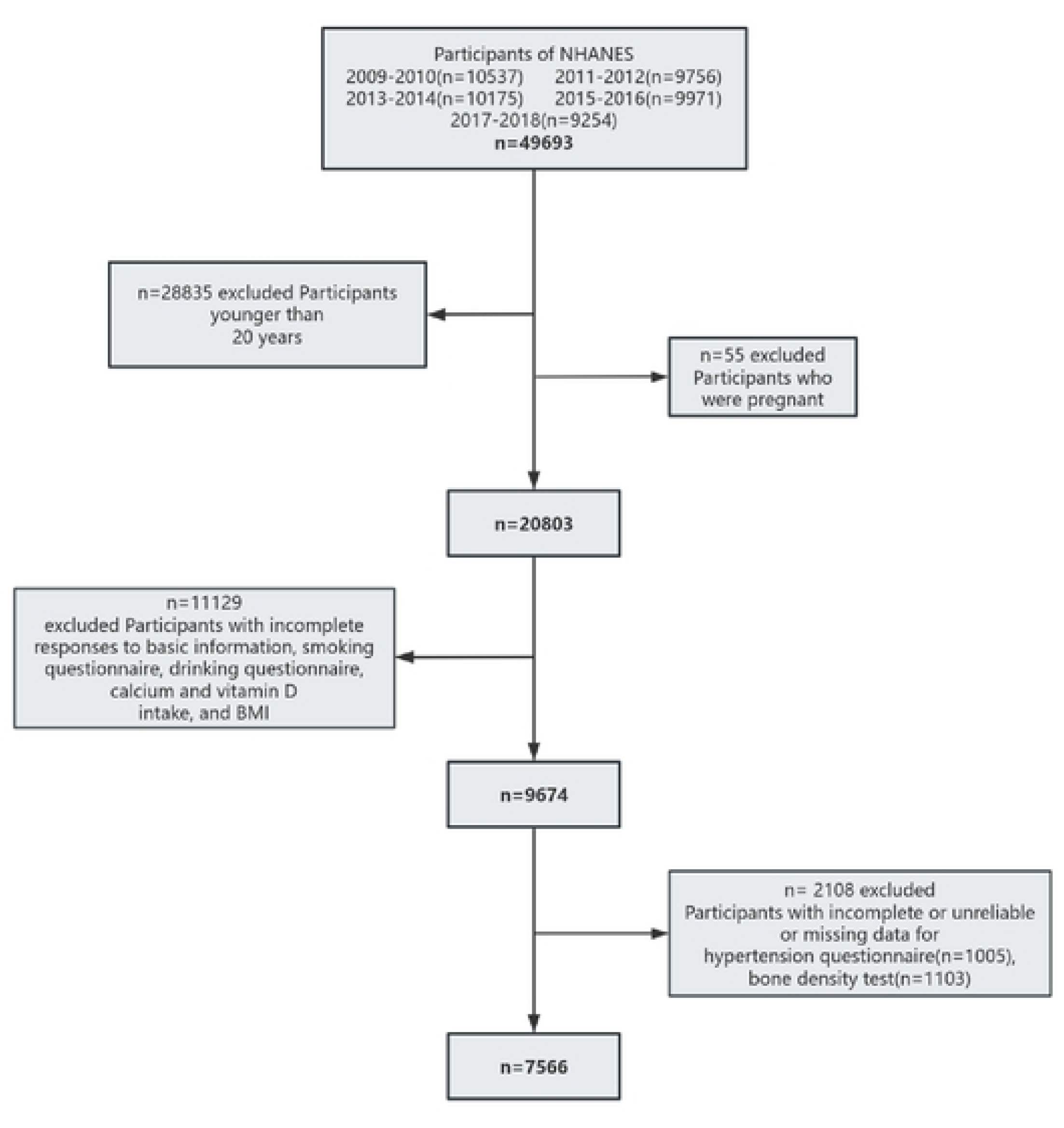
Flowchart of the selection process for eligible participants fro1n NHANES 2009-2018.

### 1.3 Definition of OP Based on BMD

The operational definition of OP is based on the T-score of BMD, which is defined as 2.5 standard deviations or more below the average value for young adult women[23]. The following formula is used to calculate the T-score: T-score = (BMD participant - BMD mean) / SD mean where BMD participant represents the observed BMD of the participant, BMD mean is the average BMD value for healthy young adults, and SD mean is the standard deviation of BMD among healthy young adults.OP is defined as a T-score ≤ −2.5, low bone mass is defined as a T-score < −1.0 and > −2.5, and T-score ≥ −1 is considered normal healthy bone mass[24]. In young adults aged 20-30 years, the mean value and standard deviation of total body BMD are 1.12 ± 0.10 g/cm² for men and 1.05 ± 0.09 g/cm² for women[25]. The T-score is specifically calculated using SPSS by applying the T-score formula with the mean BMD value and the mean SD value. The resulting T-scores are then used to classify the 7,566 samples into low bone mass and normal groups. After organizing the data, a baseline analysis is conducted.

### 1.4 Definition of Variables

Based on the NHANES database, HTN data were derived from self-reported variables in the survey section and the use of HTN medications. Bone metabolism study data were based on the dual-energy X-ray absorptiometry (DXA) total body scan data from the physical examination section. According to self-reported demographic survey data, race/ethnicity was divided into five groups: non-Hispanic white, non-Hispanic black, Mexican American, other Hispanic, and other races (including multiracial). Socioeconomic factors were defined, including access to healthcare, health insurance coverage, education level, or employment status. Education level was divided into four categories: less than high school, high school graduate or equivalent, some college or associate degree, and college graduate or above. Household income was divided into 14 categories, ranging from less than $4,999 to more than $75,000. Pregnancy status was defined as whether or not pregnant (yes or no). Vitamin D and calcium intake were measured as continuous variables to reflect the specific daily intake values of individuals. Body mass index (BMI) was also measured as a continuous variable, reflecting the ratio of an individual’s weight to height. Smoking and drinking status were categorized based on self-reports as whether or not engaged in smoking and drinking behaviors (yes or no).

### 1.5 Establishing a Model for Predicting Total Body BMD Using DXA

Since the NHANES database did not collect dual-energy X-ray absorptiometry (DXA) data for the whole body in 2009-2010, but only collected DXA data for the femur and spine, we developed a model to predict total body BMD using femur BMD and spine BMD.Existing research has also demonstrated the feasibility of this approach. For example, Looker et al. (1998) developed a regression model to predict total body BMD using femoral neck and lumbar spine BMD.They found significant correlations between these regional BMD measurements and total body BMD, resulting in the following predictive equation: Total body BMD = α + β1 × femoral BMD + β2 × spine BMD[26]. Wang et al. (2003) developed a predictive equation for total body BMD based on regional BMD measurements in children and adolescents. Their regression analysis indicated that femur and lumbar spine BMD significantly predict total body BMD, leading to the following model:Total body BMD = γ + δ₁ × Femur BMD + δ₂ × Spine BMD[27]. Yu et al. (2004) focused on Chinese women, developing a predictive equation for total body BMD from regional BMD measurements of the femoral neck and lumbar spine. Their multiple regression technique yielded a highly accurate model suitable for clinical application[28]. The resulting equation is: Total body BMD = a + b × femoral BMD + c × spine BMD. To obtain specific abc regression coefficients, we conducted regression analysis using SPSS for the 2013-2014 femur, spine, and total body DXA data, yielding the following regression coefficients(Figure 2):

**Figure 2.**
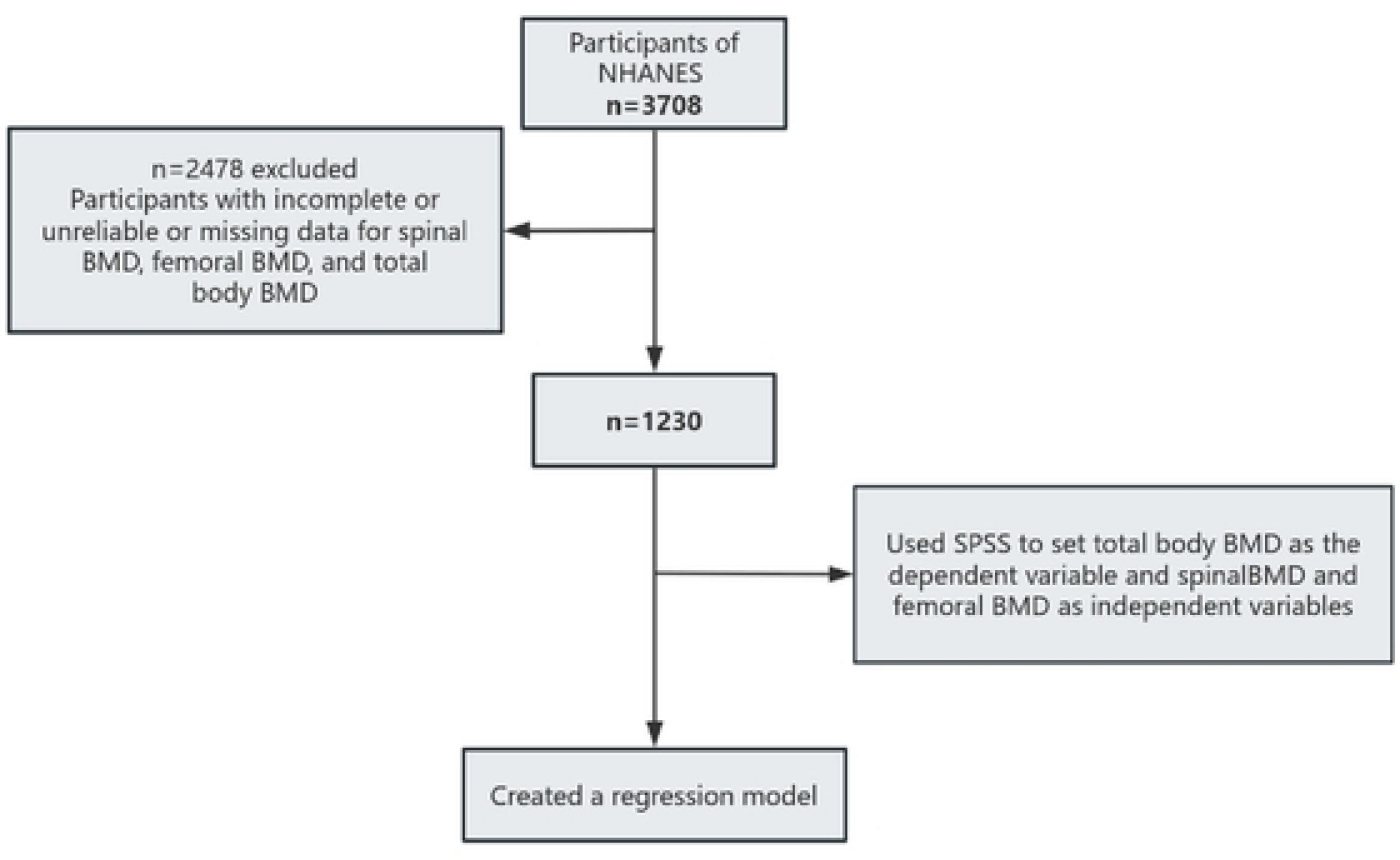
Flowchart for creating the linear regression model.

Table 1 presents the predictive results of the regression model for total BMD. The model significantly predicted total BMD, with a significant correlation between the predictor variables, femur BMD and spine BMD, and total BMD (F = 976.946, *p* < 0.001). This indicates that regression coefficients can be further calculated to quantify the specific impact of these two predictor variables on total BMD, supporting the feasibility of using femur and spine BMD to accurately estimate total BMD.

**Table 1.** ANOVA Regression Model.

| Model | Sum of Squares | df | Mean Square | Significance |
| --- | --- | --- | --- | --- |
| Regression | 9.685 | 2 | 4.842 | <.001 |
| Residual | 6.082 | 1227 | .005 |  |
| Total | 15.766 | 1229 |  |  |

**Table 2.** Coefficients of Each Variable in the Regression Analysis Unstandardized Coefficients Standardized Coefficients.

| Model | Unstandardized Coefficients |  | Standardized Coefficients |  |
| --- | --- | --- | --- | --- |
|  | B | Std. Error | Beta | Significance |
| (Constant) | .428 | .016 |  | <.001 |
| Spine BMD | .344 | .019 | .447 | <.001 |
| Femoral BMD | .332 | .020 | .404 | <.001 |

The regression equation obtained from the multivariate regression analysis of the relationship between femoral BMD, spine BMD, and total body BMD is as follows: Total body BMD = 0.428 + 0.332 × femoral BMD + 0.344 × spine BMD. The parameter estimates of the regression equation are: intercept 0.428, femoral BMD regression coefficient 0.332, and spine BMD regression coefficient 0.344. This indicates that femoral BMD and spine BMD have significant predictive capabilities for total body BMD, with *p*-values less than 0.001, demonstrating high statistical significance. By substituting the 2009-2010 spine BMD and femoral BMD data into this regression equation, total body BMD values can be predicted. This method provides a reliable basis for data cleaning and subsequent analysis of total body BMD for 2009-2010.

### 1.6 Statistical Analysis

SPSS 27.0 was used for data analysis in this study. Descriptive statistics were used to describe the basic characteristics of the sample. Continuous variables were described using means and standard deviations, and categorical variables were described using frequencies and percentages. Group comparisons were conducted using chi-square tests and multiple linear regression analysis. Multiple logistic regression analysis was used to adjust for confounding factors (such as age, gender, BMI, smoking, and drinking) to assess the independent association between HTN and OP. Finally, subgroup analyses were conducted to validate the robustness and consistency of the study results. The significance level was set at *P* < 0.05.

## Results

### 2.1 Sample Characteristics

A total of 1,477 HTN patients and 6,089 non-HTN patients were included in this study. The mean age (± standard deviation) of the HTN group was 44.37±12.548 years, while the mean age of the non-HTN group was 34.94±10.834 years, with statistically significant differences between the two groups (*p* < 0.001). In terms of gender distribution, males accounted for 76.8% of the HTN group and 60.7% of the non-HTN group (*p* < 0.001). In females, the HTN group accounted for 23.2%, and the non-HTN group accounted for 39.3% (*p* < 0.001). In terms of race/ethnicity, Mexican Americans accounted for 12.1% of the HTN group and 16.9% of the non-HTN group (*p* < 0.001); non-Hispanic whites accounted for 42.3% of the HTN group and 40.6% of the non-HTN group (*p* < 0.001); non-Hispanic blacks accounted for 25.5% of the HTN group and 17.7% of the non-HTN group (p < 0.001); and other races accounted for 20.2% of the HTN group and 24.8% of the non-HTN group (*p* < 0.001). There was no significant difference in overall education level between the HTN and non-HTN groups (*p* = 0.547), but there were significant differences in the distribution of specific education levels. For example, the proportion of the HTN group with some college education or an associate degree was 35.4%, while the non-HTN group was 33.0% (*p* = 0.003); the proportion of the HTN group with a bachelor’s degree or higher was 24.8%, while the non-HTN group was 29.4% (*p* < 0.001). There was no significant difference in annual household income between the two groups (*p* > 0.05). In terms of BMI, the mean BMI of the HTN group was 31.2082±6.66754, while the non-HTN group was 27.77±6.23048, with statistically significant differences between the two groups (*p* < 0.001). The smoking status showed that 54.4% of the HTN group were smokers, compared to 41.2% of the non-HTN group (*p* < 0.001). There was no significant difference in drinking status between the two groups (*p* = 0.106). In terms of calcium intake, the mean intake of the HTN group was 1012.84±642.236 mg, while the non-HTN group was 1019.19±620.538 mg, with no statistically significant difference between the two groups (*p* = 0.187). In terms of vitamin D intake, the mean intake of the HTN group was 4.6898±5.66105 µg, while the non-HTN group was 4.7934±5.63396 µg, with a statistically significant difference between the two groups (*p* = 0.031) (Table 3).

**Table 3.** Baseline characteristics ofHTN and non-HTN groups.

| Characteristics | Hypertension | Non-hypertension | p value |
| --- | --- | --- | --- |
| N* | 1477 | 6089 |  |
| Age† | 44.37(12.548) | 34.94(10.834) | <.001 |
| Gender |  |  | <.001 |
| Men | 1134(76.8) | 7398(60.7) |  |
| Women | 343(23.2) | 4780(39.3) |  |
| Race/Ethnicity |  |  | <.001 |
| Mexican Americans | 178(12.1) | 2058(16.9) |  |
| Other Hispanic | 125(8.5) | 1140(9.4) |  |
| Non-Hispanic White | 625(42.3) | 4950(40.6) |  |
| Non-Hispanic Black | 376(25.5) | 2160(17.7) |  |
| Other Race - Including Multi-Racial | 173(11.7) | 1870(15.4) |  |
| Education Level |  |  | .547 |
| Less than 9th grade | 67(4.5) | 632(5.2) |  |
| 9-11th grade (Includes 12th grade with no diploma) | 183(12.4) | 1356(11.1) |  |
| High school graduate/GED or equivalent | 337(22.8) | 2592(21.3) |  |
| Some college or AA degree | 523(35.4) | 4016(33) |  |
| College graduate or above | 367(24.8) | 3582(29.4) |  |
| Annual Household Income |  |  | .102 |
| \$ 0 to \$ 4,999 | 34(2.3) | 278(2.3) | |
| \$ 5,000 to \$ 9,999 | 52(3.5) | 292(2.4) | |
| \$10,000 to \$ 14,999 | 86(5.8) | 540(4.4) | |
| \$ 15,000 to \$ 19,999 | 84(5.7) | 628(5.2) | |
| \$ 20,000 to \$ 24,999 | 96(6.5) | 766(6.3) | |
| \$ 25,000 to \$ 34,999 | 134(9.1) | 1368(11.2) | |
| \$ 35,000 to \$ 44,999 | 153(10.4) | 1238(10.2) | |
| \$ 45,000 to \$ 54,999 | 118(8) | 1024(8.4) | |
| \$ 55,000 to \$ 64,999 | 87(5.9) | 828(6.8) | |
| \$ 65,000 to \$ 74,999 | 75(5.1) | 662(5.4) | |
| \$20,000 and Over | 56(3.8) | 440(3.6) | |
| Under \$20,000 | 11(0.7) | 104(0.9) | |
| \$75,000 to \$99,999 | 168(11.4) | 1402(11.5) | |
| \$100,000 and Over | 323(21.9) | 2608(21.4) | |
| BMI† | 31.2082(6.66754) | 27.77(6.23048) | <.001 |
| Smoking |  |  | <.001 |
| yes | 804(54.4) | 5012(41.2) |  |
| no | 673(45.6) | 7166(58.8) |  |
| Drinking† | 3.13(2.497) | 3.1(2.45) | .106 |
| Calcium Intake† | 1012.84(642.236) | 1019.19(620.538) | .187 |
| Vitamin D Intake† | 4.6898(5.66105) | 4.7934(5.63396) | .031 |
| BMD† | 1.1507(0.10653) | 1.1271(0.09781) | <.001 |

Note: BMI: Body Mass Index. BMD: Bone Mineral Density. p-value: Statistical significance. \*N represents unweighted numbers, and other values are weighted using NHANES MEC examination weights. †Figures are expressed as mean (standard error) for means (e.g., age, BMI), and other figures are expressed as number (percentage).

This study included a total of 266 patients with low bone mass and 7,300 patients with normal bone mass. The mean age (± standard deviation) of the low bone mass group was 34.5±11.865 years, while that of the normal bone mass group was 36.02±11.393 years, with a statistically significant difference between the two groups (*p* < 0.001). In terms of gender distribution, males accounted for 41.8% in the low bone mass group and 63.3% in the normal bone mass group (*p* < 0.001); among females, the low bone mass group accounted for 58.2% and the normal bone mass group accounted for 36.7% (*p* < 0.001). Regarding race/ethnicity, among Mexican Americans, the low bone mass group accounted for 21.2% and the normal bone mass group accounted for 16.2% (*p* < 0.001); among other Hispanics, the low bone mass group accounted for 11% and the normal bone mass group accounted for 9.2% (*p* < 0.001); among non-Hispanic whites, the low bone mass group accounted for 37.8% and the normal bone mass group accounted for 40.9% (*p* < 0.001); among non-Hispanic blacks, the low bone mass group accounted for 4.9% and the normal bone mass group accounted for 19.1% (*p* < 0.001); among other races (including multiracial), the low bone mass group accounted for 25.1% and the normal bone mass group accounted for 14.6% (*p* < 0.001). There was no significant overall difference in education level between the low bone mass and normal bone mass groups (*p* = 0.531), but there were significant differences in the distribution of specific education levels. For example, the proportion of the low bone mass group with some college education or an associate degree was 35.7%, while that of the normal bone mass group was 33.1%; the proportion of the low bone mass group with a bachelor’s degree or higher was 29.6%, while that of the normal bone mass group was 28.9%. In terms of annual household income, there was no statistically significant difference between the two groups (*p* > 0.05). The mean BMI of the low bone mass group was 25.2769±6.09462, while that of the normal bone mass group was 28.2485±6.35445, with a statistically significant difference (*p* < 0.001). Smoking status showed that 38% of the low bone mass group were smokers, compared to 42.8% of the normal bone mass group (*p* = 0.059). There was no significant difference in drinking status between the two groups (*p* = 0.176). In terms of calcium intake, the mean intake of the low bone mass group was 904.94±556.528 mg, while that of the normal bone mass group was 1022.73±624.856 mg, with a statistically significant difference (*p* < 0.001). Regarding vitamin D intake, the mean intake of the low bone mass group was 3.9947±5.25681 µg, while that of the normal bone mass group was 4.8115±5.64849 µg, with no statistically significant difference (*p* = 0.989). Regarding HTN, 8.6% of the low bone mass group had HTN, compared to 10.9% of the normal bone mass group (*p* = 0.006) (Table 4).

**Table 4.**
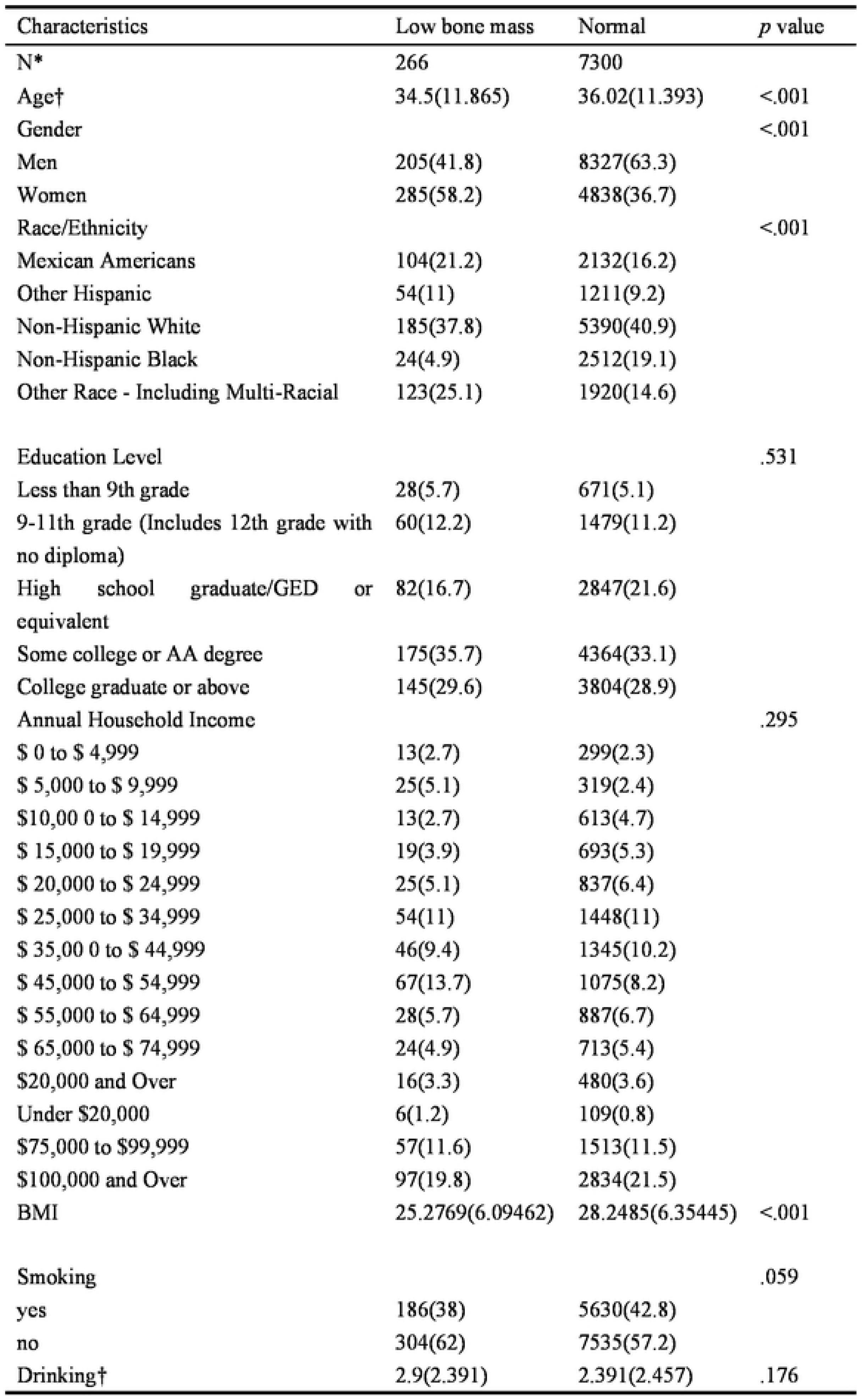
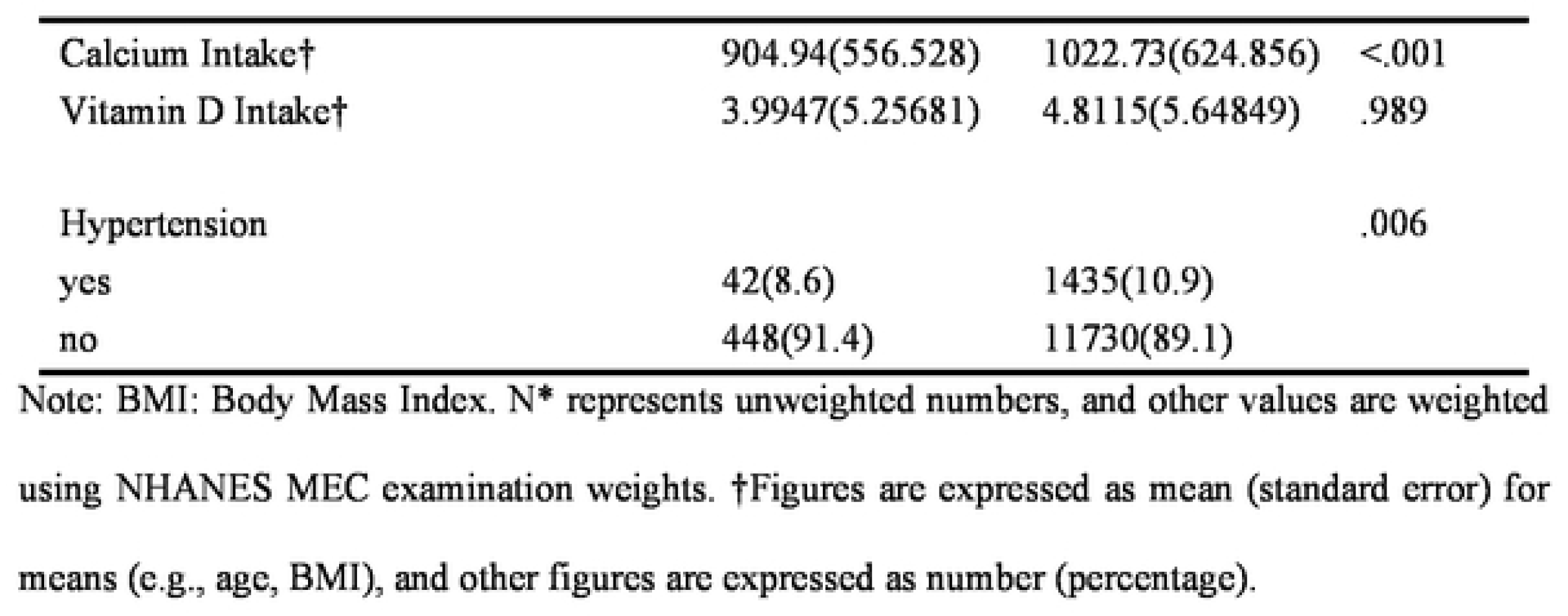
Baseline Characteristics of Patients with Low Bone Mass and Normal Bone Mass.

### 2.2 Relationship Between HTN and Bone Metabolism Abnormalities

The correlation analysis between HTN and BMD, both before and after adjusting for covariates, shows that HTN is significantly associated with BMD [Beta = −0.024 (95% CI: −0.029 to −0.018), *p* < 0.001]. This association remained significant after adjusting for age, gender, race, and BMI [Beta = −0.007 (95% CI: −0.013 to −0.002), *p* = 0.007]. Further adjustments for socioeconomic factors (including education level and household income) still showed a significant association [Beta = −0.007 (95% CI: - 0.013 to −0.002), *p* = 0.006]. After adjusting for vitamin D and calcium intake, the association remained significant [Beta = −0.007 (95% CI: −0.013 to −0.002), *p* = 0.007]. Finally, after adjusting for smoking and alcohol consumption, the association continued to be significant [Beta = −0.007 (95% CI: −0.013 to −0.002), *p* = 0.006] (Table 5).

**Table 5.** Correlation Between HTN and BMD.

|  | Beta (95% CI) | *p*-value |
| --- | --- | --- |
| Crude | -0.024(-0.029--0.018) | <.001 |
| Model 1 | -0.007(-0.013--0.002) | 0.007 |
| Model 2 | -0.007(-0.013--0.002) | 0.006 |
| Model 3 | -0.007(-0.013--0.002) | 0.007 |
| Model 4 | -0.007(-0.013--0.002) | 0.006 |

Note: Values are expressed as regression coefficients (95% confidence interval). Model 1: Adjusted for age, gender, race, and BMI. Model 2: Further adjusted for socioeconomic factors, including education level and household income. Model 3: Further adjusted for vitamin D and calcium intake. Model 4: Further adjusted for smoking and alcohol consumption.

### 2.3 Subgroup Analysis

For subgroup analysis, the study recoded age (a continuous variable) into three age groups: 20-39 years, 40-59 years, and 60 years and above. Using the “Recode into Different Variables” function in SPSS, the age variable was recoded by range (20-39, 40-59, 60 and above) into a new categorical variable “Age Group”. The new age grouping variable was then used in subsequent stratified regression analyses to explore the relationship between HTN and BMD across different age groups.

The subgroup analysis of the association between HTN and BMD shows that HTN remains significantly associated with BMD after adjustment [Beta = −0.007 (95% CI: - 0.013 to −0.002), *p* = 0.006]. Gender-stratified analysis reveals a significant negative correlation between HTN and BMD in men [Beta = −0.01 (95% CI: −0.016 to −0.003), *p* = 0.004], while the association is not significant in women [Beta = −0.006 (95% CI: - 0.015 to 0.004), *p* = 0.244]. Age-stratified analysis shows that HTN is significantly negatively correlated with BMD in the 40-59 age group [Beta = −0.01 (95% CI: −0.017 to −0.002), *p* = 0.012], while the association is not significant in the 20-39 age group [Beta = −0.006 (95% CI: −0.014 to 0.002), *p* = 0.131] and in those aged 60 and above [Beta = −0.001 (95% CI: −0.023 to 0.021), *p* = 0.946] (Table 6).

**Table 6.**
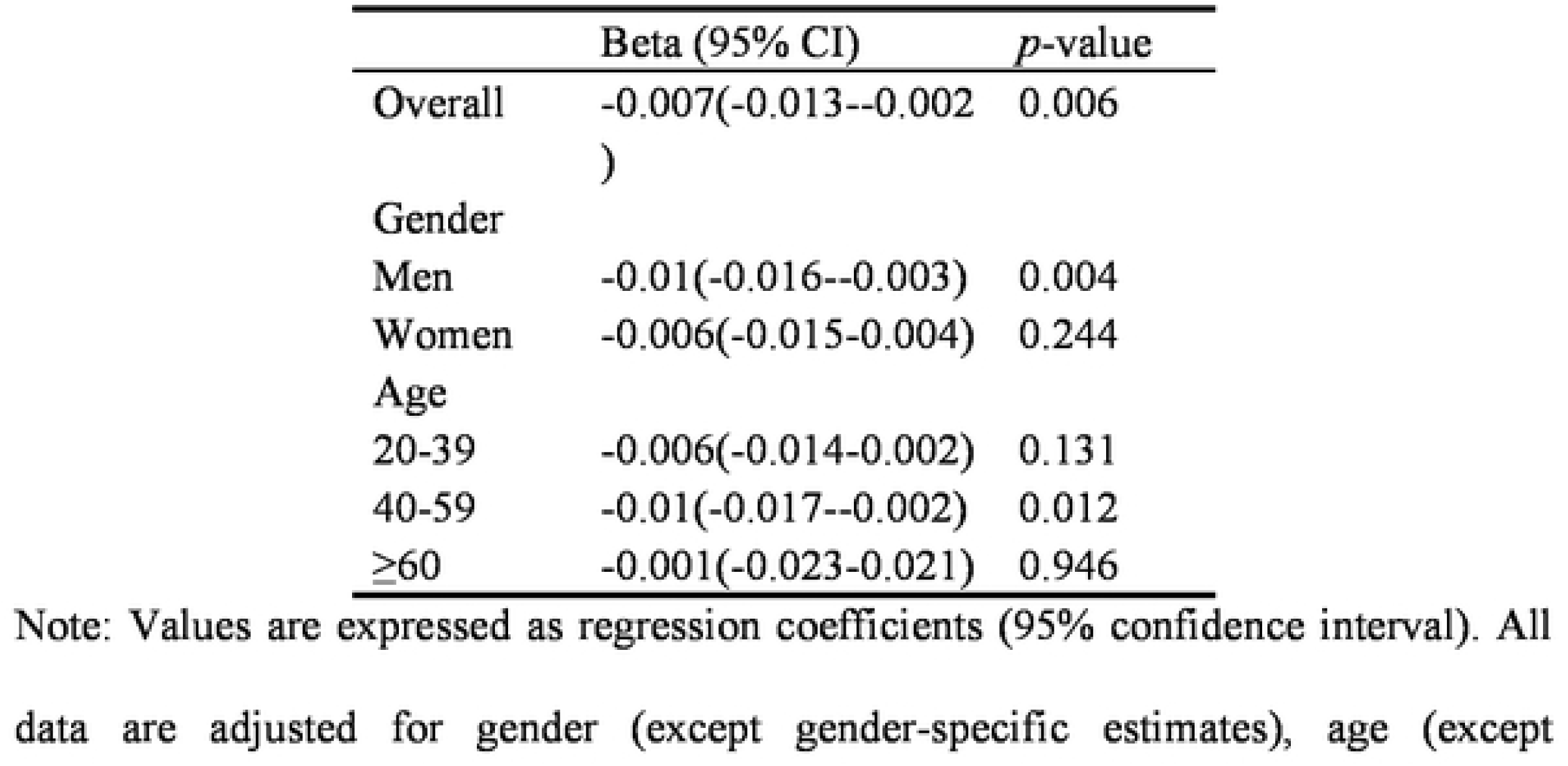
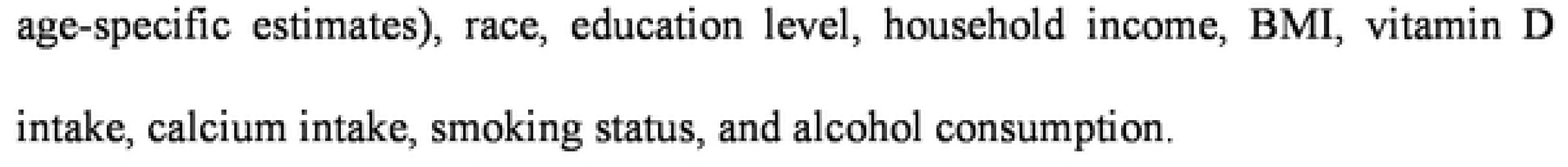
Subgroup Analysis of the Correlation Between HTN and BMD.

|  | Beta (95% CI) | *p*-value |
| --- | --- | --- |
| Overall | -0.007(-0.013--0.002) | 0.006 |
| Gender |  |  |
| Men | -0.01(-0.016--0.003) | 0.004 |
| Women | -0.006(-0.015-0.004) | 0.244 |
| Age |  |  |
| 20-39 | -0.006(-0.014-0.002) | 0.131 |
| 40-59 | -0.01(-0.017--0.002) | 0.012 |
| ≥60 | -0.001(-0.023-0.021) | 0.946 |

Note: Values are expressed as regression coefficients (95% confidence interval). All data are adjusted for gender (except gender-specific estimates), age (except
age-specific estimates), race, education level, household income, BMI, vitamin D intake, calcium intake, smoking status, and alcohol consumption.

### 2.4 Constructing Predictive Models with SHAP

SHAP value analysis revealed that although the correlation between HTN (BAQ0) and BMD (BMXBMI) is small, it remains statistically significant. This indicates that while HTN is not a primary determinant of BMD changes, it does have some influence on the model’s predictive outcomes. In addition to HTN, other features such as RIDRETH1 (race/ethnicity), RIAGENDR (gender), and BMXBMI (body mass index) also show significant influence in both graphs, with BMXBMI playing a particularly important role in predicting BMD (Figure 3A). After cross-validation and hyperparameter optimization, the machine learning model’s decision boundary is shown (Figure 3B). The optimized model may have made more precise adjustments in the distribution of feature importance, providing more accurate predictions.

**Figure 3.**
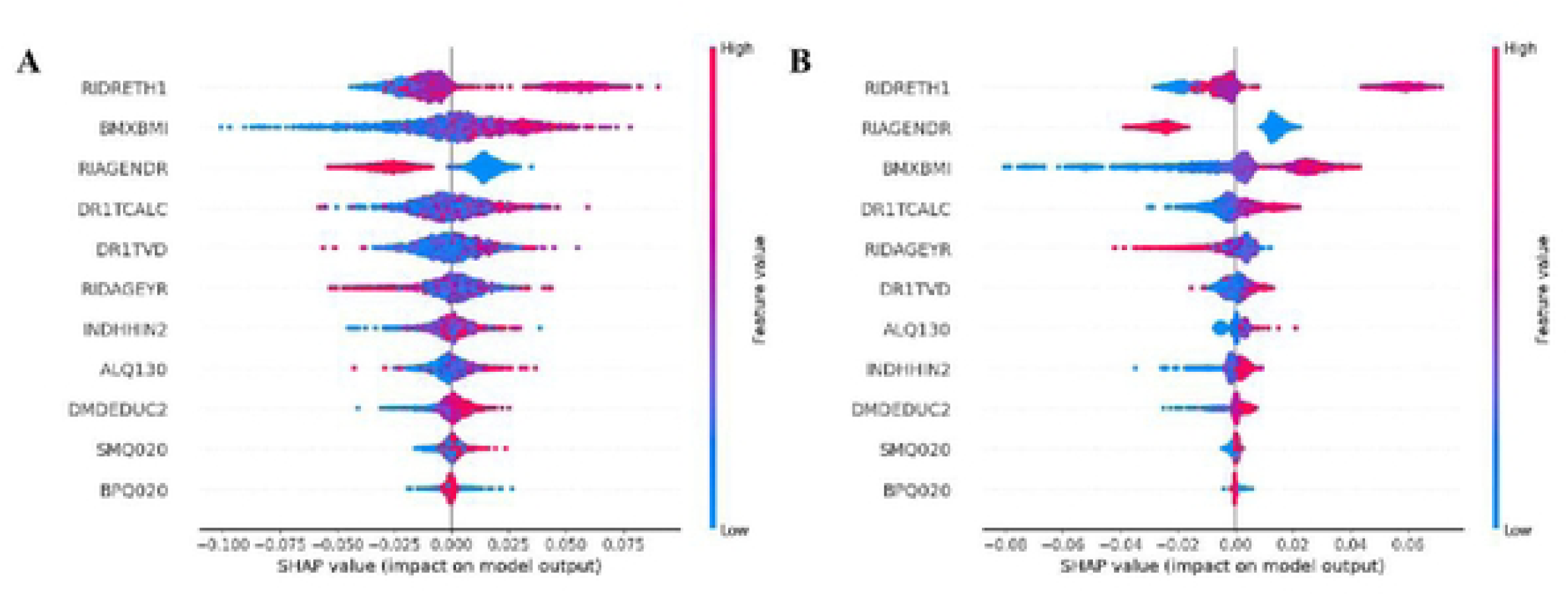
A, SHAP Model Prediction Correlation. B, SHAP Cross-Validation and Hyperparameter Optimization Model. Note: Figure 3 illustrates the feature importance and their impact on the output in predictive models using various machine learning algorithms (such as linear regression, decision tree, random forest, XGBoost, etc.). This figure is generated based on SHAP (SHapley Additive exPlanations) values, with each dot representing a feature’s impact on the model output. The position shows the magnitude of the SHAP value, and the color ranges from blue to red, indicating the change in feature values from low lo high. The mean squared error (MSE) of the neural network model is 0.0090, the decision tree model’s MSE is 0.0166, the random forest model’s MSE is 0.0087, and the XGBoost model’s MSE is 0.0091.

## Discussion

HTN and OP are major public health issues, and there may be common metabolic pathways between them[29]. The NHANES database, with its multistage probability sampling design, provides a nationally representative sample covering demographic information, health status, and nutritional status, ensuring the comprehensiveness and reliability of the study results[30]. Through continuous data accumulation over the years, NHANES supports trend analysis and long-term health monitoring[31]. This study combines traditional statistical analysis with machine learning models, specifically using SHAP value analysis to reveal the subtle relationship between HTN and BMD.. Although the correlation is small, the application of machine learning models enhances the stability and interpretability of the results, providing strong support for understanding complex medical phenomena[32]. Compared to previous studies that were limited by insufficient sample sizes, lack of data diversity, and the use of singular analytical methods, this study fully leverages long-term trend data for in-depth analysis. Additionally, it makes extensive use of modern machine learning techniques to enhance the stability and interpretability of the results[33].This study introduces innovative methods to estimate total body BMD and further validates these methods using machine learning algorithms, with the aim of providing a more comprehensive and detailed understanding of the relationship between HTN and OP. Initially, the study utilizes the NHANES database for traditional SPSS-based analysis. The NHANES database, known for its extensive and diverse dataset, supports large-scale analyses, thereby enhancing the representativeness and generalizability of the findings. By integrating advanced machine learning algorithms, the study delves deeper into the data, offering a more nuanced analysis. The application of machine learning techniques not only facilitates the handling of large-scale and high-dimensional data but also reveals hidden patterns and complex relationships that traditional statistical methods might overlook, thereby significantly enhancing the accuracy and reliability of the data analysis. Furthermore, the combination of traditional statistical methods with machine learning approaches offers a novel perspective on the relationship between HTN and OP, addressing the limitations of previous studies that relied solely on conventional data analysis. This integrative approach markedly improves the interpretability and predictive power of the findings. It not only aids in uncovering potential pathological mechanisms underlying the relationship between HTN and OP, thereby providing more detailed and reliable scientific evidence, but also offers new insights for clinical practice. The study suggests that in the management of hypertensive patients, close attention should be paid to BMD monitoring and osteoporosis prevention. This could lead to the development of more individualized prevention and treatment strategies, ultimately improving patient quality of life and reducing the overall disease burden, which holds significant implications for both clinical practice and public health.

### Hypertension and Bone Mass: Comparative Analysis

This study identified significant differences between the HTN and non-HTN groups through descriptive statistical analysis. Compared to the non-HTN group, the HTN group exhibited a higher average age, a greater proportion of males, higher BMI, and a higher smoking rate. However, no significant differences were found between the two groups in terms of education level, household income, alcohol consumption, and intake of calcium and vitamin D. Furthermore, notable differences were observed between the low bone mass group and the normal bone mass group regarding age, gender, race/ethnicity, BMI, calcium intake, and HTN status. Specifically, low bone mass was significantly associated with younger age, a higher proportion of females, a greater prevalence of non-Hispanic whites, lower BMI, and HTN. Additionally, calcium intake was significantly correlated with BMD, while factors such as education level, household income, smoking, and alcohol consumption exhibited weaker associations with BMD and did not reach statistical significance.

### Analysis of the Impact of Hypertension on Bone Health

The study indicates a significant association between the prevalence of HTN and OP in the general US population, independent of traditional confounding factors. These findings suggest that HTN patients should be screened for OP, and corresponding preventive strategies should be formulated based on different populations to alleviate the burden of OP. Moreover, HTN may affect bone metabolism through various mechanisms, increasing the risk of OP and fractures[34,35]. HTN can lead to vascular dysfunction, affecting blood flow to the bones, which in turn impacts the nutrition and metabolism of bone cells. These hemodynamic changes may result in a decrease in BMD[36]. HTN patients often experience abnormal calcium metabolism, such as increased urinary calcium excretion and decreased serum calcium levels. This calcium metabolism imbalance weakens the bone mineralization process, making bones more fragile[37]. For OP patients with a history of fractures, HTN may increase calcium loss in the urine, leading to a negative calcium balance in bone remodeling and further reducing BMD[38]. HTN is also closely related to chronic low-grade inflammation, which increases levels of pro-inflammatory cytokines that can affect bone metabolism by disrupting bone formation and promoting bone resorption[39]. HTN may also affect the levels of various hormones in the body, including parathyroid hormone and cortisol[40–42], which directly impact bone metabolism, while elevated cortisol may increase fracture risk by inhibiting bone formation[43]. Epidemiological studies have found that as blood pressure increases, the rate of mineral loss from bones accelerates, further decreasing BMD in OP patients with a history of fractures[44]. HTN is associated with high concentrations of parathyroid hormone, which can accelerate bone turnover, leading to bone loss[45]. HTN may impair brain structures related to gait control and balance, leading to falls and subsequent fractures[46]. Additionally, medications used to treat HTN, such as diuretics and certain antihypertensive drugs, may also indirectly affect BMD by influencing calcium metabolism and excretion[47]. Diuretics can increase urinary calcium excretion, thereby lowering calcium levels in the body, affecting bone health[48]. Therefore, HTN management should not only focus on cardiovascular health but also protect bone health. Combining current research progress, this study further explores the epidemiological evidence of the association between HTN and bone metabolism among NHANES participants. The results indicate that after fully adjusting for confounding factors, a significant association remains between HTN and bone metabolism. From a clinical perspective, this finding is important. First, HTN may impact bone health by increasing the risk of OP and fractures, especially in patients with poorly controlled HTN[49]. Therefore, monitoring BMD becomes a necessary measure in the management of patients with HTN. Although the impact of HTN on BMD is relatively small, its potential adverse effects should not be overlooked. This necessitates the consideration of bone health factors when developing HTN treatment plans.Additionally, the study suggests strengthening bone health protection strategies in the comprehensive management of HTN patients to reduce fracture risk. These findings provide valuable evidence for clinical practice, emphasizing the importance of considering BMD in the treatment of HTN.

### Limitations

Although this study confirmed the correlation between HTN and OP, there are several limitations that need to be discussed. Initially, although this study explored the relationship between femur BMD and spine BMD with total body BMD through regression analysis and successfully estimated total body BMD for the 2009-2010 period, some limitations still exist.The first limitation is that the characteristics of the sample and environmental factors at different time periods may vary, potentially affecting the accuracy and external validity of the results. Second, socioeconomic status, healthcare quality, and lifestyle factors may change over different years, and these changes could potentially impact the BMD measurements.Third, the regression model assumes a stable linear relationship between variables, but the actual situation may be more complex. Thus, the regression coefficients derived from 2013-2014 data applied to 2009-2010 data may have certain biases and limitations. Furthermore, although weighted data was used to improve the representativeness of the analysis, the generalizability of the results may be affected by the accuracy and applicability of the weights used, given that the data was sourced from an epidemiological survey. Therefore, the conclusions of the study may not fully represent the actual situation of the entire U.S. population. Lastly, while machine learning algorithms provided a deeper understanding of the relationship between HTN and bone metabolism, the results of these models are limited by data quality and algorithm selection. Specifically, the predictive performance and explanatory power of machine learning models depend on the feature selection of the input data and the tuning of model parameters, and issues such as model complexity and overfitting may affect the stability and generalization ability of the results. Therefore, future research should focus on strengthening the collection of long-term follow-up data to verify the potential causal relationship between HTN and bone metabolism abnormalities. Additionally, it is necessary to further explore the specific biological mechanisms and intervention strategies to apply new therapeutic methods in clinical practice and mitigate the negative impact of HTN on bone health.

## Conclusion

In conclusion, this study reveals a significant negative correlation between HTN and bone metabolism abnormalities, emphasizing the importance of focusing on bone health in HTN patients. The study results support incorporating BMD monitoring into HTN management, and future efforts should focus on developing effective prevention strategies to reduce the risk of OP.

## Data Availability

All data produced in the present study are available upon reasonable request to the authors
All data produced in the present work are contained in the manuscript
All data produced are available online at

https://www.cdc.gov/nchs/nhanes/index.htm

## Author Contributions

Conceptualization: Jinyao Li, Mingcong Tang, Ziqi Deng, Yanchen Feng, Xue Dang, Lu Sun, Yunke Zhang, Jianping Yao, Min Zhao, Feixiang Liu

Data curation: Jinyao Li, Mingcong Tang, Ziqi Deng

Formal analysis: Jinyao Li, Mingcong Tang, Ziqi Deng, Yanchen Feng, Xue Dang, Lu Sun, Feixiang Liu

Funding acquisition: Yunke Zhang, Jianping Yao, Min Zha

Feixiang Liu Investigation: Jinyao Li, Mingcong Tang, Ziqi Deng, Yanchen Feng, Xue Dang, Lu Su

Feixiang Liu Methodology: Jinyao Li, Mingcong Tang, Ziqi Deng, Yanchen Feng, Xue Dang, Lu Sun

Project administration: Yunke Zhang, Jianping Yao, Min Zhao, Feixiang Liu Software: Jinyao Li

Supervision: Jinyao Li, Mingcong Tang, Ziqi Deng, Yanchen Feng, Xue Dang, Lu Su

Feixiang Liu Validation: Jinyao Li, Mingcong Tang, Ziqi Deng, Yanchen Feng, Xue Dang, Lu Sun, Yunke Zhang, Jianping Yao, Min Zha Feixiang Liu

Visualization: Jinyao Li

Writing– original draft: Jinyao Li, Mingcong Tang, Ziqi Deng, Yanchen Feng, Xue Dang, Lu Sun

Writing– review & editing: Jinyao Li, Mingcong Tang, Ziqi Deng, Yanchen Feng, Xue Dang, Lu Sun, Yunke Zhang, Jianping Yao, Min Zhao, Feixiang Liu

## References

1. Burnier M, Damianaki A. Hypertension as cardiovascular risk factor in chronic kidney disease. Circulation research. 2023 Apr 14;132(8):1050–63.

2. World Health Organization. Hypertension [Internet]. 2023 Mar 16 [cited 2024 Aug 6]. Available from: https://www.who.int/news-room/fact-sheets/detail/hypertension.

3. Chapman N, Ching SM, Konradi AO, Nuyt AM, Khan T, Twumasi-Ankrah B, Cho EJ, Schutte AE, Touyz RM, Steckelings UM, Brewster LM. Arterial hypertension in women: state of the art and knowledge gaps. Hypertension. 2023 Jun;80(6):1140–9.

4. Song S, Guo Y, Yang Y, Fu D. Advances in pathogenesis and therapeutic strategies for osteoporosis. Pharmacology & therapeutics. 2022 Sep 1;237:108168.

5. Barnsley J, Buckland G, Chan PE, Ong A, Ramos AS, Baxter M, Laskou F, Dennison EM, Cooper C, Patel HP. Pathophysiology and treatment of osteoporosis: challenges for clinical practice in older people. Aging clinical and experimental research. 2021 Apr;33:759–73.

6. Huang Y, Ye J. Association between hypertension and osteoporosis: a population-based cross-sectional study. BMC Musculoskeletal Disorders. 2024 Jun 3;25(1):434.

7. Matei A, Bilha SC, Constantinescu D, Pavel-Tanasa M, Cianga P, Covic A, Branisteanu DD. Body composition, adipokines, FGF23-Klotho and bone in kidney transplantation: Is there a link?. Journal of nephrology. 2021 Feb 9:1–2.

8. Zhivodernikov IV, Kirichenko TV, Markina YV, Postnov AY, Markin AM. Molecular and cellular mechanisms of osteoporosis. International journal of molecular sciences. 2023 Oct 30;24(21):15772.

9. Ciancia S, van Rijn RR, Högler W, Appelman-Dijkstra NM, Boot AM, Sas TC, Renes JS. Osteoporosis in children and adolescents: when to suspect and how to diagnose it. European journal of pediatrics. 2022 Jul;181(7):2549–61.

10. Zhang W, Luo Y, Xu J, Guo C, Shi J, Li L, Sun X, Kong Q. The possible role of electrical stimulation in osteoporosis: A narrative review. Medicina. 2023 Jan 8;59(1):121.

11. Mo L, Ma C, Wang Z, Li J, He W, Niu W, Chen Z, Zhou C, Liu Y. Integrated bioinformatic analysis of the shared molecular mechanisms between osteoporosis and atherosclerosis. Frontiers in Endocrinology. 2022 Jul 22;13:950030.

12. Xu J, Yu L, Liu F, Wan L, Deng Z. The effect of cytokines on osteoblasts and osteoclasts in bone remodeling in osteoporosis: A review. Frontiers in Immunology. 2023 Jul 5;14:1222129.

13. Liu X, Liu X, Wang Y, Zeng B, Zhu B, Dai F. Association between depression and oxidative balance score: National Health and Nutrition Examination Survey (NHANES) 2005–2018. Journal of affective disorders. 2023 Sep 15;337:57–65.

14. Younossi ZM, Paik JM, Al Shabeeb R, Golabi P, Younossi I, Henry L. Are there outcome differences between NAFLD and metabolic-associated fatty liver disease?. Hepatology. 2022 Nov;76(5):1423–37.

15. Mahemuti N, Jing X, Zhang N, Liu C, Li C, Cui Z, Liu Y, Chen J. Association between systemic immunity-inflammation index and hyperlipidemia: a population-based study from the NHANES (2015–2020). Nutrients. 2023 Feb 26;15(5):1177.

16. Huang Q, Wan J, Nan W, Li S, He B, Peng Z. Association between manganese exposure in heavy metals mixtures and the prevalence of sarcopenia in US adults from NHANES 2011–2018. Journal of hazardous materials. 2024 Feb 15;464:133005.

17. Safaei M, Sundararajan EA, Driss M, Boulila W, Shapi’i A. A systematic literature review on obesity: Understanding the causes & consequences of obesity and reviewing various machine learning approaches used to predict obesity. Computers in biology and medicine. 2021 Sep 1;136:104754.

18. Theodosiou AA, Read RC. Artificial intelligence, machine learning and deep learning: Potential resources for the infection clinician. Journal of Infection. 2023 Jul 17.

19. Lello S, Capozzi A, Scambia G. Osteoporosis and cardiovascular disease: an update. Gynecological Endocrinology. 2015 Aug 3;31(8):590–4.

20. Matsumoto T, Sone T, Soen S, Tanaka S, Yamashita A, Inoue T. Abaloparatide increases lumbar spine and hip BMD in Japanese patients with osteoporosis: the phase 3 ACTIVE-J study. The Journal of Clinical Endocrinology & Metabolism. 2022 Oct 1;107(10):e4222–31.

21. Xing W, Gao W, Zhao Z, Xu X, Bu H, Su H, Mao G, Chen J. Dietary flavonoids intake contributes to delay biological aging process: analysis from NHANES dataset. Journal of Translational Medicine. 2023 Jul 21;21(1):492.

22. Looker AC, Wahner HW, Dunn WL, Calvo MS, Harris TB, Heyse SP, Johnston Jr CC, Lindsay R. Updated data on proximal femur bone mineral levels of US adults. Osteoporosis international. 1998 Aug;8:468–90.

23. Xue S, Zhang Y, Qiao W, Zhao Q, Guo D, Li B, Shen X, Feng L, Huang F, Wang N, Oumer KS. An updated reference for calculating bone mineral density T-scores. The Journal of Clinical Endocrinology & Metabolism. 2021 Jul 1;106(7):e2613–21.

24. Bouffard J, Weber Z, Pearsall L, Emery K, Côté JN. Similar effects of fatigue induced by a repetitive pointing task on local and remote light touch and pain perception in men and women. Plos one. 2020 Dec 18;15(12):e0244321.

25. Uddin MN, Li LZ, Deng BY, Ye J. Interpretable XGBoost–SHAP machine learning technique to predict the compressive strength of environment-friendly rice husk ash concrete. Innovative Infrastructure Solutions. 2023 May;8(5):147.

26. Zanker CL, Gannon L, Cooke CB, Gee KL, Oldroyd B, Truscott JG. Differences in bone density, body composition, physical activity, and diet between child gymnasts and untrained children 7-8 years of age. Journal of bone and mineral research. 2003 Jun;18(6):1043–50.

27. Yu W, Glüer CC, Fuerst T, Grampp S, Li J, Lu Y, Genant HK. Influence of degenerative joint disease on spinal bone mineral measurements in postmenopausal women. Calcified Tissue International. 1995 Sep;57:169–74.

28. Evenepoel P, Cunningham J, Ferrari S, Haarhaus M, Javaid MK, Lafage-Proust MH, Prieto-Alhambra D, Torres PU, Cannata-Andia J. European Consensus Statement on the diagnosis and management of osteoporosis in chronic kidney disease stages G4–G5D. Nephrology Dialysis Transplantation. 2021 Jan;36(1):42–59.

29. Mejía Sandoval HJ. Prevalencia de osteoporosis posmenopáusica de muy alto riesgo en el Hospital Universitario Nacional.

30. Wu WT, Li YJ, Feng AZ, Li L, Huang T, Xu AD, Lyu J. Data mining in clinical big data: the frequently used databases, steps, and methodological models. Military Medical Research. 2021 Dec;8:1–2.

31. Dye BA, Afful J, Thornton-Evans G, Iafolla T. Overview and quality assurance for the oral health component of the National Health and Nutrition Examination Survey (NHANES), 2011–2014. BMC Oral Health. 2019 Dec;19:1–1.

32. Kim JI, Maguire F, Tsang KK, Gouliouris T, Peacock SJ, McAllister TA, McArthur AG, Beiko RG. Machine learning for antimicrobial resistance prediction: current practice, limitations, and clinical perspective. Clinical microbiology reviews. 2022 Sep 21;35(3):e00179–21.

33. Liu S, Wu S, Bao X, Ji J, Ye Y, Guo J, Liu J, Wang X, Zhang Y, Hao D, Huang D. Changes in Blood Pressure is Associated with Bone Loss in US Adults: A Cross-Sectional Study from NHANES 2005–2018. Calcified Tissue International. 2024 Mar;114(3):276–85.

34. Rodríguez-Carrio J, Martínez-Zapico A, Cabezas-Rodríguez I, Benavente L, Pérez-Álvarez ÁI, López P, Cannata-Andía JB, Naves-Díaz M, Suárez A. Clinical and subclinical cardiovascular disease in female SLE patients: interplay between body mass index and bone mineral density. Nutrition, Metabolism and Cardiovascular Diseases. 2019 Feb 1;29(2):135–43.

35. Zhang Y, Zhao C, Zhang H, Chen M, Meng Y, Pan Y, Zhuang Q, Zhao M. Association between serum soluble α-klotho and bone mineral density (BMD) in middle-aged and older adults in the United States: a population-based cross-sectional study. Aging Clinical and Experimental Research. 2023 Oct;35(10):2039–49.

36. du Toit WL, Kruger R, Gafane-Matemane LF, Schutte AE, Louw R, Mels CM. Urinary metabolomics profiling by cardiovascular risk factors in young adults: the African Prospective study on Early Detection and Identification of Cardiovascular disease and Hypertension study. Journal of Hypertension. 2022 Aug 1;40(8):1545–55.

37. Suh B, Yu H, Kim H, Lee S, Kong S, Kim JW, Choi J. Interpretable deep-learning approaches for osteoporosis risk screening and individualized feature analysis using large population-based data: Model development and performance evaluation. Journal of medical Internet research. 2023 Jan 13;25:e40179.

38. Quereda C, Orte L, Sabater J, Navarro-Antolin J, Villafruela JJ, Ortuno J. Urinary calcium excretion in treated and untreated essential hypertension. Journal of the American Society of Nephrology. 1996 Jul 1;7(7):1058–65.

39. Foessl I, Dimai HP, Obermayer-Pietsch B. Long-term and sequential treatment for osteoporosis. Nature Reviews Endocrinology. 2023 Sep;19(9):520–33.

40. Cho HW, Jin HS, Eom YB. FGFRL1 and FGF genes are associated with height, hypertension, and osteoporosis. Plos one. 2022 Aug 18;17(8):e0273237.

41. Brown JP. Long-term treatment of postmenopausal osteoporosis. Endocrinology and Metabolism. 2021 Jun 22;36(3):544–52.

42. Yu B, Wang CY. Osteoporosis and periodontal diseases–An update on their association and mechanistic links. Periodontology 2000. 2022 Jun;89(1):99–113.

43. Pan M, Pan X, Zhou J, Wang J, Qi Q, Wang L. Update on hormone therapy for the management of postmenopausal women. Bioscience trends. 2022 Feb 28;16(1):46–57.

44. Hu Z, Yang K, Hu Z, Li M, Wei H, Tang Z, Chen B, Su C, Cai D, Xu J. Determining the association between hypertension and bone metabolism markers in osteoporotic patients. Medicine. 2021 Jun 18;100(24):e26276.

45. Cappuccio FP, Kalaitzidis R, Duneclift S, Eastwood JB. Unravelling the links between calcium excretion, salt intake, hypertension, kidney stones and bone metabolism. Journal of nephrology. 2000 May 1;13(3):169–77.

46. Bromfield SG, Ngameni CA, Colantonio LD, Bowling CB, Shimbo D, Reynolds K, Safford MM, Banach M, Toth PP, Muntner P. Blood pressure, antihypertensive polypharmacy, frailty, and risk for serious fall injuries among older treated adults with hypertension. Hypertension. 2017 Aug;70(2):259–66.

47. Wu D, Cline-Smith A, Shashkova E, Perla A, Katyal A, Aurora R. T-cell mediated inflammation in postmenopausal osteoporosis. Frontiers in immunology. 2021 Jun 30;12:687551.

48. Xiao PL, Cui AY, Hsu CJ, Peng R, Jiang N, Xu XH, Ma YG, Liu D, Lu HD. Global, regional prevalence, and risk factors of osteoporosis according to the World Health Organization diagnostic criteria: a systematic review and meta-analysis. Osteoporosis International. 2022 Oct;33(10):2137–53.

49. Ayers C, Kansagara D, Lazur B, Fu R, Kwon A, Harrod C. Effectiveness and safety of treatments to prevent fractures in people with low bone mass or primary osteoporosis: a living systematic review and network meta-analysis for the American College of Physicians. Annals of internal medicine. 2023 Feb;176(2):182–95.

